# Building Large-Scale Registries from Unstructured Clinical Notes using a Low-Resource Natural Language Processing Pipeline

**DOI:** 10.1101/2022.12.23.22283914

**Authors:** Nazgol Tavabi, James Pruneski, Shahriar Golchin, Mallika Singh, Ryan Sanborn, Benton Heyworth, Amir Kimia, Ata Kiapour

## Abstract

Building clinical registries is an important step in improving the quality and safety of patient care. With the growing size of medical records, manual abstraction becomes more and more infeasible and impractical. On the other hand, Natural Language Processing Techniques have shown promising results in extracting valuable information from unstructured clinical notes. However, the structure and nature of clinical notes are very different from regular text that state-of-the-art NLP models are trained and tested on and they have their own set of challenges. In this study, we propose SE-K, an efficient and interpretable classification approach for extracting information from clinical notes, and show that it outperforms current state-of-the-art models in text classification. We use this approach to generate a 20-year comprehensive registry of anterior cruciate ligament reconstruction operations, one of the most common orthopedics operations among children and young adults. This registry can help us better understand the outcomes of this surgery and identify potential areas for improvement which can ultimately lead to better treatment outcomes.

## 1 Introduction

Medical records have long been an essential source of information for improving the quality of patient care [1, 2]. Electronic health records (EHR) consist of structured and unstructured data containing laboratory reports, medical history, reports of operative interventions, and other medical services provided to the patient. Most often, researchers rely on large datasets to perform clinical research, and these cohorts are commonly identified by structured EHR data, specifically current procedural terminology (CPT) codes for operations and procedures. In addition to structured EHR data, manual abstraction from the medical record has traditionally been used to extract patient information for biomedical studies. This method, however, is both expensive and labor-intensive, and can pose risks to patient privacy. Additionally, structured EHR data, including diagnostic codes, has been shown to capture some conditions unreliably [3–5].

Given the rapidly increasing amount of unstructured data, there is increasing interest in using natural language processing (NLP) and machine learning (ML) in registry building as an alternative to traditional methods (e.g., manual chart reviews). Notably, up to 80% of the electronic health record consists of unstructured data, providing an opportunity for NLP to become an invaluable tool to automate the processing and characterization of clinical texts into cohort-specific registries [6–10]. Additionally, automating the process of analyzing clinical notes and building registries can help reduce human error [7].

Over the past years, several information-extraction (IE) approaches have been developed and adapted to build high-quality databases to automate building large registries from clinical notes which otherwise would not have been possible with manual chart review [11–15]. In this paper we propose two approaches for extracting information from clinical notes and use them to develop a 20-year comprehensive registry of anterior cruciate ligament (ACL) reconstruction operations, one of the most common orthopedics operations among children and young adults. This registry is built based on high volume data from a pediatric and young-adult orthopedic and sports medicine (OSM) academic practice. We compare the results from the proposed approaches with Bidirectional Encoder Representations from Transformers [16] (BERT, current State-Of-The-Art in NLP).

## 2 Methods

### 2.1 Data

Following IRB approval (IRB-P00037878), notes from patients with at least one encounter at any of the six Boston’s Children Hospital Orthopedic Surgery and Sports Medicine clinics from 2000 – 2021 were acquired. ACL surgery CPT code (CPT-29888) was used to train a model to identify arthroscopically aided anterior cruciate ligament repair/augmentation or reconstruction surgeries from operative notes, based on a previously described approach,[7] with an area under the receiver operating characteristic curve (AUROC) of 0.99 and accuracy of 1. This was done to address the common mislabelling and missing lable issues associated with cohort identification based on procedure codes.

Once these notes were acquired, they were used to extract patient-specific details of the ACL procedures to build a registry of all the operations performed in Boston’s Children Hospital Orthopedic Surgery and Sports Medicine clinics. The variables extracted from operative notes and their descriptions are shown in Table 1.

**Table 1:** Variables extracted from operative notes.

| Variable | Description | Options |
| --- | --- | --- |
| Laterality | Surgical Side | Left<br>Right |
| Revision | Revision ACL Surgery | First<br>Revision |
| Graft Type | ACL Graft Type | Auto-Hams<br>Auto-BTB<br>Auto-itb<br>Allograft<br>Auto-quad<br>BEAR<br>Hams+Allo |
| Interference Screw | Interference Screw used for graft fixation | True<br>False |
| Endobutton | Endobutton used for graft fixation | True<br>False |
| Tightrope | Tightrope use for reconstruction | True<br>False |
| Medial Meniscus | Medial Meniscus Surgery | Repair<br>Meniscectomy<br>None |
| Lateral Meniscus | Lateral Meniscus Surgery | Repair<br>Meniscectomy<br>None |

Additionally, some variables of interest for the registry were described in clinical notes prior to the operation (pre-operative notes). To extract these variables, orthopedic clinic notes within 180 days of the surgery were acquired for each of the patients identified by the model and classified as pre-operative notes. The variables extracted from pre-operative notes are shown in Table 2.

**Table 2:** Variables extracted from pre-operative notes.

| Variable | Description | Options |
| --- | --- | --- |
| Laterality | Surgical Side | Left<br>Right<br>Unknown |
| Inj Mech | Injury Mechanism | True<br>False |
| Tear Type | Type of ACL Tear | Complete<br>Partial<br>Sprain<br>Unknown |
| Skeletal Age | Skeletal Maturity Status | Mature<br>Immature<br>Unknown |
| Ipsi ACL reinjury | Ipsilateral ACL Reinjury | True<br>False |
| MCL Injury | Concomitant MCL Injury | True<br>False |
| LatMen Injury | Concomitant Lateral Meniscus Injury | True<br>False |
| MedMen Injury | Concomitant Medial Meniscus Injury | True<br>False |
| PCL Injury | Concomitant PCL Injury | True<br>False |
| PLC Injury | Concomitant PLC Injury | True<br>False |

A subset of clinical notes obtained within 2000-2020 and all the data from 2021 were manually labeled using a custom developed graphic user interface DrT (Document review Tool,) [17, 18] to reduce the time required for human review of charts with a number of key user interface features. The manual labeling was done for variables in Tables 1 and 2 and used for training and testing the models. We held the 2021 data for validation.

### 2.2 Sentence Extractor with Keywords

Our first approach is Sentence Extractor with Keywords (SE-K). This approach is based on Term Frequency–Inverse Document Frequency (TF-IDF), which is a statistical method for extracting features from text [19]. First TF-IDF features are obtained from the clinical notes. Then K most important keywords are identified using F-values from ANOVA test on TF-IDF features, where K is a hyper-parameter and the search space for K is set as log-uniform (in Hyperopt [20]) so that the model has a higher chance of assigning a lower number to K to avoid overfitting. Figure 3 shows important keywords identified for each variable.

Based on the most important keywords, sentences in the note are ranked such that sentences with more important keywords are ranked higher. The sentences are ranked based on the inner products of F-values and TF-IDF values of the most important keywords. Then *K′* number of the most important sentences from each note are extracted. *K′* is another hyper-parameter with uniform distribution where 0 < K′ ⩽ 10. The inner product of the chosen sentences has to be non-zero, meaning that if the note has less than *K′* sentences with important keywords, the model would output fewer sentences.

Features of the important sentences are first concatenated to the features of the whole note before using the filter to remove features with a correlation higher than 95%, then *K* most relevant features are chosen and fed to the classifier. A graphical representation of this pipeline is shown in Figure 1.

**Fig 1:**
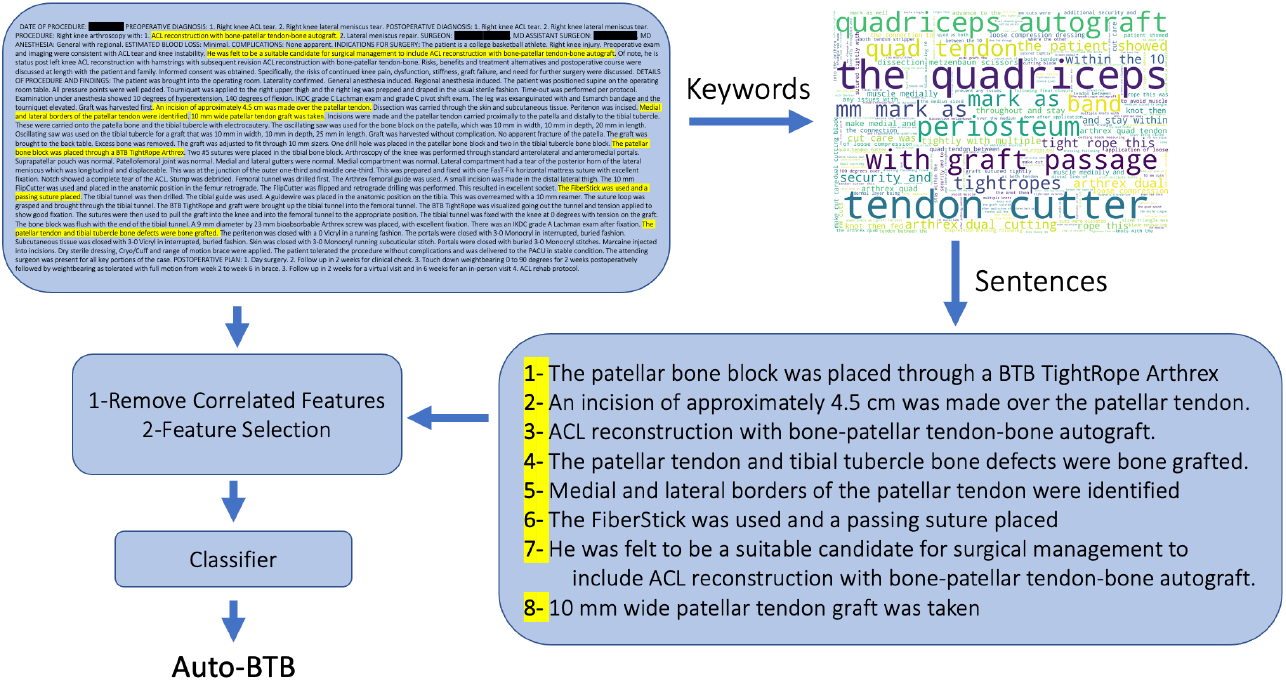
An overview of SE-K pipeline. First, important keywords are extracted from clinical notes. Then, important sentences are identified based on the keywords. Features from both the whole text and the important sentences are extracted and used for classification. In this example, the target variable is “Graft Type”. (The highlighted sentences in the whole note are the important sentences shown in the bottom right graph.)

### 2.3 Sentence Extractor with Embeddings

Our second approach is Sentence Extractor with Embeddings (SE-E). This approach is similar to SE-K mentioned previously in Section 2.2, the difference is in the representation of the *K′* selected sentences.

With SE-E, in addition to TF-IDF representations, doc2vec [21] representations of the important sentences are also provided for classification. Text embedding methods have been shown to improve representations by taking the context of the text into account compared to keyword-based methods like TF-IDF and Bag-Of-Words [22, 23]. Doc2vec [21] utilizes a shallow network for generating sentence-level embeddings and unlike most other embedding methods it does not need GPUs or large datasets to train, hence, it is used here to extract embeddings of important sentences.

### 2.4 BERT

BERT [16] is a State-of-the-Art NLP model which has been extensively used in the healthcare domain [7, 24–26]. The self-attention mechanisms inherent to this model calculate the relationships between all pairs of tokens, allowing the model to identify the most informative words and phrases in each document. Because of the model’s large architecture, it requires a substantial amount of training data and in most cases, the pre-trained versions of these models are used. In this work we used Bio-Clinical BERT [27, 28], which has been pretrained on PubMed Corpus [29] and publicly available MIMIC dataset [30, 31] and we further trained it on 23,871,108 clinical notes from Boston Children’s Hospital (BCH BERT)[24].

An issue with using BERT for the tasks in this study is that the input to the model should be less than 512 tokens, if it is longer the model truncates the text meaning it ignores the rest of the text. However, most clinical notes are longer than 512 tokens and the variable of interest might be discussed near the end of the note.

### 2.5 Majority Vote

Finally, we used a hard voting approach, [32, 33] which is an ensemble method, based on the three sets of predictions available from SE-K, SE-E, and BERT. Ensemble methods have been used to improve results in numerous studies [34–36]. In most cases ensemble methods have the best performance since they take advantage of the results of all other approaches, on the other hand, they are least efficient in terms of time and computation resources [37, 38].

### 2.6 Comparison and Analysis

For each variable and approach, a model was trained with the same 80%-20% train-test split of manually labeled data (2000-2020). For SE-K and SE-E classification was completed using a Support Vector Machine (SVM) with Radial Basis Function kernel (RBF) [39, 40]. Hyper-parameters *C* and *γ* (SVM hyper-parameters) and *K* and *K′* were set using Hyperopt based on 5-fold validation on the train set. For BERT, 12.5% of the train set (10% of manually labeled data) was set as validation and BCH BERT was fine-tuned on the down-stream task for 10 epochs. the best model based on validation was returned and used for predicting the labels for the test set. For each variable, the AUROC, accuracy, sensitivity, and specificity were measured on the test set, for multiclass variables Average of One-vs-Rest AUROC, sensitivity, and specificity were computed. Different approaches were compared with each other based on a Critical Difference Diagram [41, 42] on AUROC of their variables. This approach is used to compare multiple classifiers over multiple data sets or problems, and it ranks the classifiers based on AUROC, where rank 1 indicates the best performing classifier, which is seen on the right of the diagram in Figures 2 and 6. The diagram denotes a lack of significant difference in AUROC by connecting similar classifiers with a thick horizontal line. This diagram is computed based on the Friedman test and a post-hoc analysis based on the Wilcoxon-Holm method. All P-values are two-sided and significant at P<0.05.

**Fig 2:**
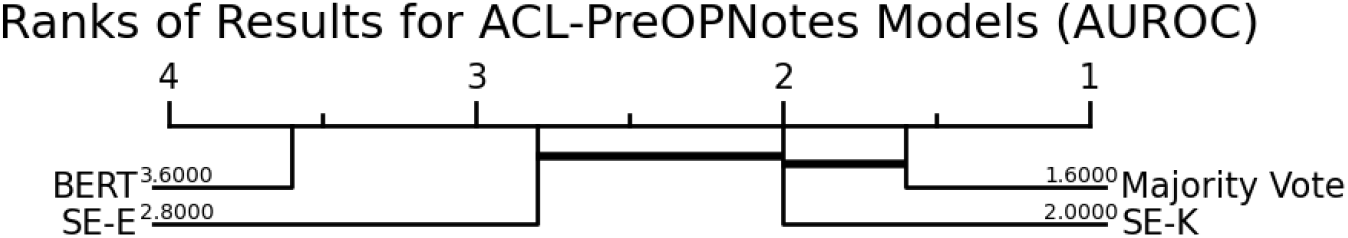
Ranking of different approaches based on AUROC in predicting the pre-operative variables. The horizontal lines indicate lack of significant difference between the 2 approach.

### 2.7 Out-of-Sample Validation

In order to evaluate the generalizability of the approaches and their ability to be used in practice, we used the models trained on data from the years 2000-2020 to predict and extract the variables of interest from notes collected in 2021.

### 2.8 Efficacy Assessment

To assess the efficacy of each approach, the running time during which the model was trained and used to predict the test set was calculated across all operative and pre-operative variables. For BERT models, only the fine-tuning time was considered and the time required for pre-training was ignored.

## 3 Results

Our model identified 5,895 operative notes detailing ACL surgeries, including 1,169 (19.8%) that were not assigned CPT code 29888 (arthroscopically aided anterior cruciate ligament repair/augmentation or reconstruction). Using the medical record numbers (MRNs) associated with identified operative reports and the time of the surgery, 9,382 pre-operative notes were identified for these patients.

### 3.1 Relative Performance on Test Data

Performance metrics for the four approaches in extracting the information from both the pre-operative and operative notes on the test set (2000-2020) are presented in Table 3. For the operative notes, there are no significant differences in AUROC between the four approaches (null hypothesis of the Friedman test over the entire classifiers cannot be rejected, P-value = 0.06). The ensemble majority vote has the highest AUROC on 7 out of 8 variables with 4 of them being tied with other approaches. SE-K is the second-best performing model, with the highest AUROC on 3 out of 8 variables with 2 of them being ties. With regards to the pre-operative variables, the majority vote has the best results in terms of AUROC followed by SE-K, while there is no significant difference between the two approaches (P=0.98). However, the majority vote has a significantly better performance than SE-E (P=0.04) and BERT (P=0.04), and SE-K has a significantly better performance than BERT (P=0.03). The ranking of approaches on pre-operative variables is shown in Figure 2. Figure 3 shows word-clouds generated by SE-K with identified important keywords for all variables in the operative and pre-operative notes, words with larger fonts have more importance in predicting the variable. Additionally, Figure 4 shows average BERT embeddings for ACL operative notes shown in 2 dimensions using PCA [43] and TSNE [44, 45].

**Table 3:** Prediction results of different approaches on pre-operative and operative variables (2000-2020 data). For each variable, the highest AUROC is highlighted in bold. In the case of ties, the highest AUROCs are highlighted in italics.

|  | SE-K |  |  |  | SE-E |  |  |  | BERT |  |  |  | Majority Vote |  |  |  |
| --- | --- | --- | --- | --- | --- | --- | --- | --- | --- | --- | --- | --- | --- | --- | --- | --- |
|  | Acc | ROC | Spec | Sen | Acc | ROC | Spec | Sen | Acc | ROC | Spec | Sen | Acc | ROC | Spec | Sen |
| Laterality | 1.0 | *1.0* | 1.0 | 1.0 | 1.0 | *1.0* | 1.0 | 1.0 | 1.0 | *1.0* | 1.0 | 1.0 | 1.0 | *1.0* | 1.0 | 1.0 |
| Revision | 0.98 | 0.97 | 0.98 | 0.96 | 0.99 | 0.93 | 0.99 | 0.88 | 1.0 | **0.98** | 1.0 | 0.96 | 1.0 | 0.97 | 1.0 | 0.94 |
| Graft Type | 0.98 | 0.98 | 0.96 | 0.99 | 1.0 | 0.99 | 0.99 | 1.0 | 1.0 | 0.99 | 0.97 | 1.0 | 1.0 | **1.0** | 0.99 | 1.0 |
| Interference Screw | 1.0 | **1.0** | 1.0 | 1.0 | 0.98 | 0.96 | 0.92 | 1.0 | 0.97 | 0.94 | 0.9 | 0.98 | 0.98 | 0.96 | 0.92 | 1.0 |
| Endobutton | 0.95 | *0.97* | 1.0 | 0.93 | 0.98 | *0.97* | 0.94 | 0.99 | 0.97 | 0.96 | 0.93 | 0.99 | 0.98 | *0.97* | 0.95 | 0.99 |
| Tightrope | 1.0 | 0.99 | 1.0 | 0.99 | 1.0 | *1.0* | 1.0 | 1.0 | 0.99 | 0.95 | 1.0 | 0.9 | 1.0 | *1.0* | 1.0 | 1.0 |
| Medial Meniscus | 0.99 | 0.97 | 0.96 | 0.98 | 0.98 | 0.95 | 0.93 | 0.97 | 0.99 | 0.97 | 0.96 | 0.98 | 0.99 | **0.98** | 0.97 | 0.99 |
| Lateral Meniscus | 0.99 | 0.98 | 0.98 | 0.99 | 0.98 | 0.97 | 0.96 | 0.98 | 0.99 | 0.99 | 0.98 | 0.99 | 0.99 | **0.99** | 0.99 | 0.99 |
| Laterality | 0.99 | *0.99* | 0.99 | 0.99 | 0.98 | 0.98 | 0.97 | 0.98 | 0.99 | 0.98 | 0.97 | 0.99 | 0.99 | *0.99* | 0.99 | 0.99 |
| Inj Mech | 0.95 | 0.95 | 0.97 | 0.92 | 0.98 | 0.97 | 0.99 | 0.95 | 0.99 | **0.99** | 0.99 | 0.99 | 0.98 | 0.97 | 0.99 | 0.95 |
| Tear Type | 0.99 | **0.96** | 0.94 | 0.98 | 0.98 | 0.9 | 0.81 | 0.98 | 0.96 | 0.81 | 0.65 | 0.96 | 0.99 | 0.91 | 0.83 | 0.99 |
| Skeletal Age | 1.0 | *0.99* | 0.98 | 1.0 | 1.0 | *0.99* | 0.99 | 1.0 | 0.99 | 0.82 | 0.76 | 0.89 | 1.0 | *0.99* | 0.99 | 1.0 |
| Ipsi ACL reinjury | 0.95 | **0.96** | 0.95 | 0.98 | 0.97 | 0.95 | 0.97 | 0.92 | 0.97 | 0.91 | 0.98 | 0.84 | 0.97 | 0.95 | 0.97 | 0.93 |
| MCL Injury | 0.99 | *0.99* | 0.98 | 0.99 | 0.98 | 0.98 | 0.98 | 0.99 | 0.91 | 0.91 | 0.94 | 0.87 | 0.99 | *0.99* | 0.99 | 0.99 |
| LatMen Injury | 0.98 | 0.98 | 0.98 | 0.99 | 0.98 | 0.98 | 0.98 | 0.99 | 0.92 | 0.92 | 0.96 | 0.88 | 0.99 | **0.99** | 0.98 | 0.99 |
| MedMen Injury | 0.98 | 0.98 | 0.98 | 0.98 | 0.98 | 0.98 | 0.98 | 0.98 | 0.91 | 0.91 | 0.92 | 0.9 | 0.99 | **0.99** | 0.98 | 0.99 |
| PCL Injury | 0.96 | 0.94 | 0.98 | 0.9 | 0.98 | *0.96* | 1.0 | 0.92 | 0.93 | 0.91 | 0.95 | 0.87 | 0.98 | *0.96* | 1.0 | 0.92 |
| PLC Injury | 0.94 | 0.94 | 0.93 | 0.96 | 0.94 | 0.93 | 0.94 | 0.93 | 0.85 | 0.86 | 0.84 | 0.87 | 0.95 | **0.96** | 0.94 | 0.97 |

**Fig 3:**
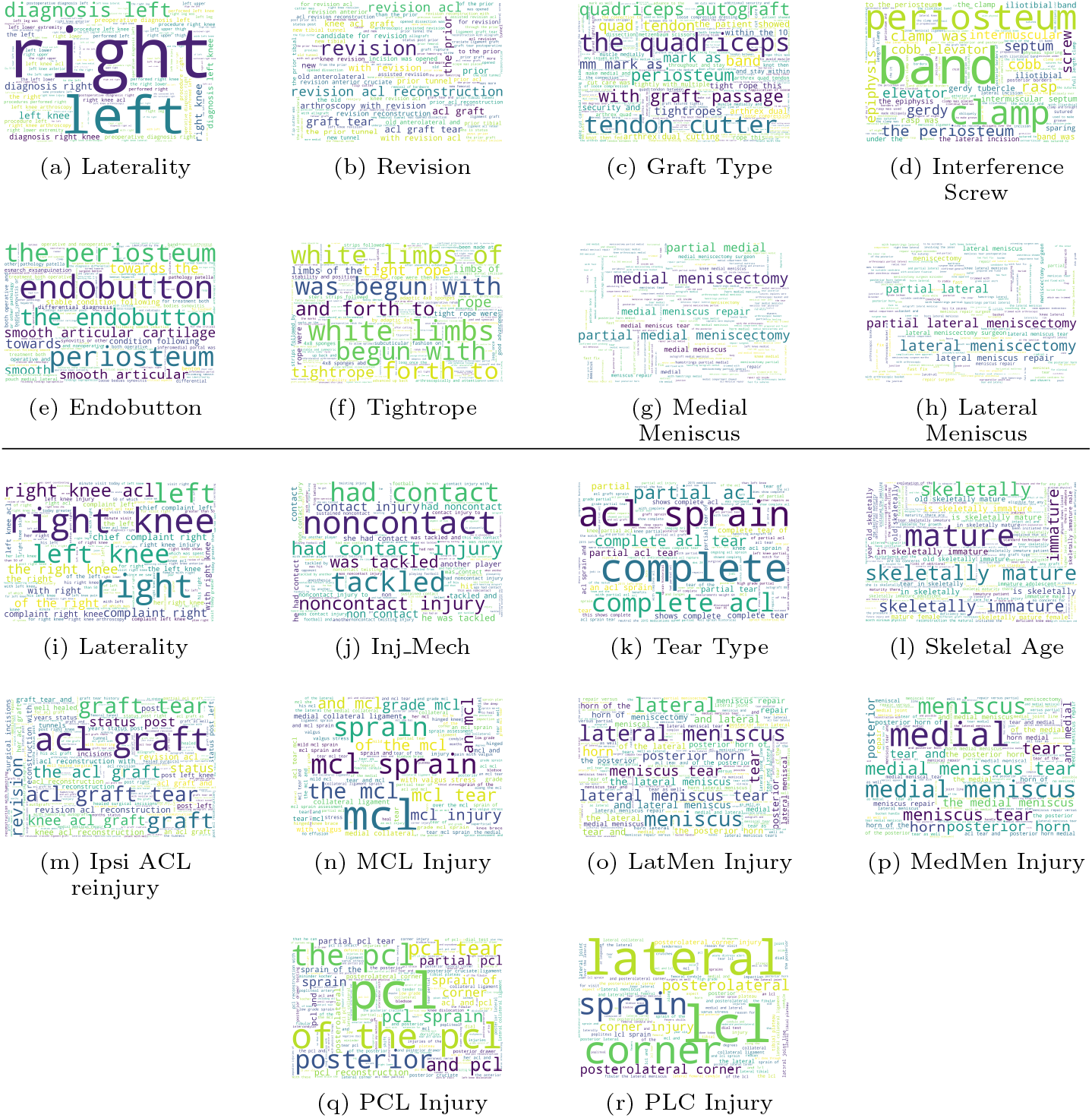
Most important keywords in predicting different variables in ACL reconstruction operative and pre-operative notes.

**Fig 4:**
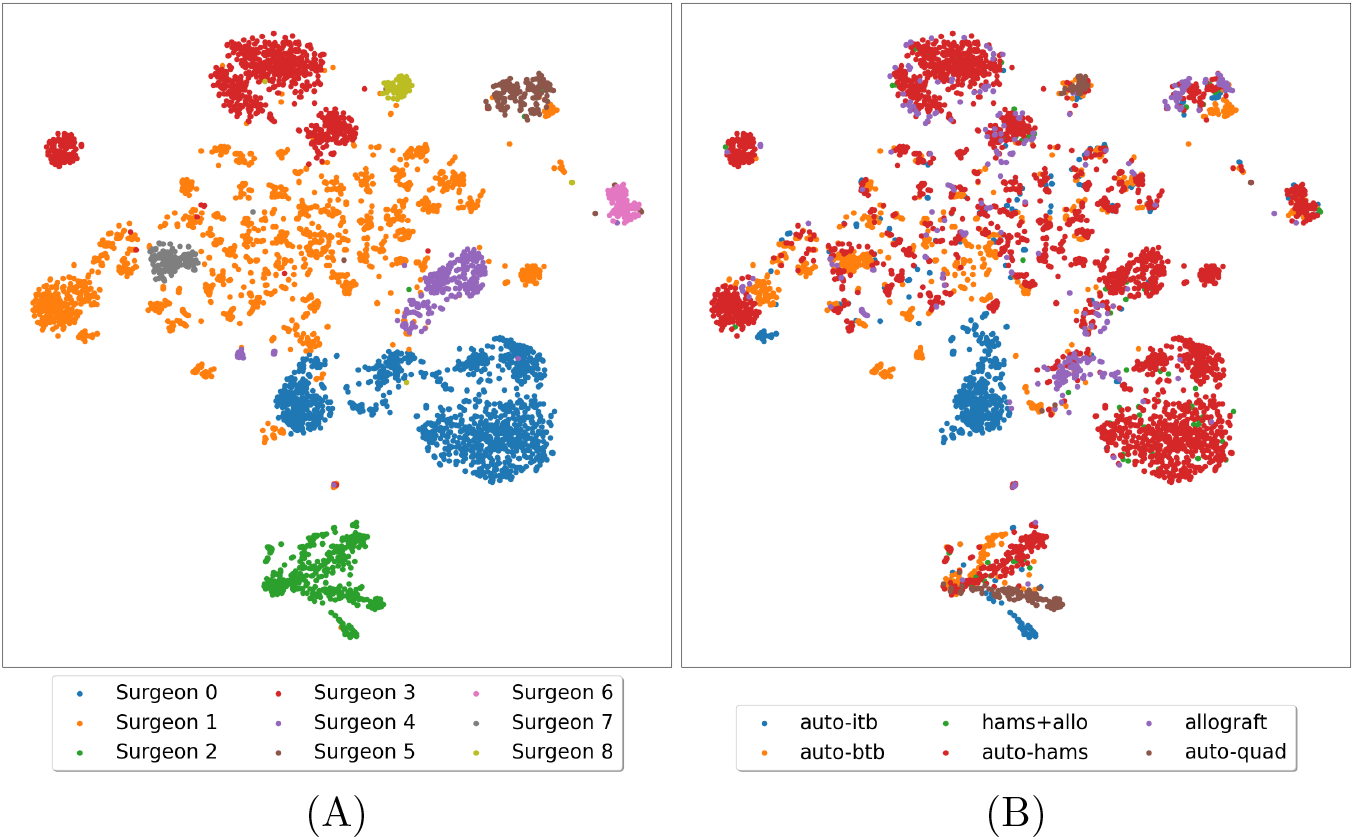
Visualizing operative notes’ BERT embeddings on 2 dimensions using T-SNE & PCA. The left plot is colored based on the primary surgeon of the operation and the right plot is based on the graft type used in the surgery which is an important factor that many other details in the surgery depend on.

### 3.2 Out-of-Sample Validation

The out-of-sample validation data is shown in Table 4. In general, there is a slight decrease in performance for all models when predicting variables from 2021 notes compared to using random test sets from 2000-2020 notes. The average change in performance for each model can be seen in Figure 5. The BERT models have a significantly larger performance decrease for the out-of-sample notes compared to SE-K (P-value=0.009) and the ensemble majority vote (P-value: 0.033), suggestive of overfitting.

**Table 4:** Prediction results of different approaches on pre-operative and operative variables on data from 2021. For each variable, the highest AUROC is highlighted in bold. In the case of ties, the highest AUROCs are highlighted in italics.

|  | SE-K |  |  |  | SE-E |  |  |  | BERT |  |  |  | Majority Vote |  |  |  |
| --- | --- | --- | --- | --- | --- | --- | --- | --- | --- | --- | --- | --- | --- | --- | --- | --- |
|  | Acc | ROC | Spec | Sen | Acc | ROC | Spec | Sen | Acc | ROC | Spec | Sen | Acc | ROC | Spec | Sen |
| Laterality | 1.0 | *1.0* | 1.0 | 1.0 | 1.0 | *1.0* | 1.0 | 1.0 | 1.0 | *1.0* | 1.0 | 1.0 | 1.0 | *1.0* | 1.0 | 1.0 |
| Revision | 0.98 | **0.95** | 0.99 | 0.91 | 0.97 | 0.91 | 0.99 | 0.83 | 0.97 | 0.91 | 0.99 | 0.83 | 0.97 | 0.91 | 0.99 | 0.83 |
| Graft Type | 0.98 | 0.96 | 0.94 | 0.99 | 1.0 | 0.99 | 0.98 | 1.0 | 1.0 | **1.0** | 1.0 | 1.0 | 1.0 | 0.99 | 0.98 | 1.0 |
| Interference Screw | 0.97 | **0.98** | 1.0 | 0.96 | 0.95 | 0.97 | 1.0 | 0.94 | 0.89 | 0.92 | 0.97 | 0.87 | 0.95 | 0.97 | 1.0 | 0.95 |
| Endobutton | 0.95 | 0.95 | 0.96 | 0.95 | 0.95 | 0.95 | 0.93 | 0.96 | 0.94 | 0.94 | 0.95 | 0.94 | 0.97 | **0.97** | 0.97 | 0.96 |
| Tightrope | 0.99 | *1.0* | 0.99 | 1.0 | 0.97 | 0.97 | 0.97 | 0.98 | 0.95 | 0.9 | 1.0 | 0.8 | 1.0 | *1.0* | 1.0 | 1.0 |
| Medial Meniscus | 0.98 | 0.93 | 0.91 | 0.96 | 0.97 | 0.93 | 0.9 | 0.96 | 0.97 | **0.95** | 0.94 | 0.96 | 0.98 | 0.93 | 0.9 | 0.96 |
| Lateral Meniscus | 0.95 | 0.94 | 0.92 | 0.96 | 0.96 | 0.94 | 0.93 | 0.96 | 0.97 | **0.97** | 0.96 | 0.98 | 0.96 | 0.95 | 0.94 | 0.96 |
| Laterality | 0.97 | *0.97* | 0.97 | 0.98 | 0.95 | 0.89 | 0.83 | 0.96 | 0.95 | 0.95 | 0.95 | 0.95 | 0.97 | *0.97* | 0.97 | 0.97 |
| Inj Mech | 0.93 | 0.88 | 0.94 | 0.83 | 0.93 | 0.92 | 0.93 | 0.9 | 0.9 | 0.88 | 0.91 | 0.86 | 0.96 | **0.93** | 0.97 | 0.9 |
| Tear Type | 0.95 | **0.92** | 0.87 | 0.97 | 0.92 | 0.83 | 0.72 | 0.94 | 0.9 | 0.74 | 0.56 | 0.91 | 0.95 | 0.87 | 0.78 | 0.96 |
| Skeletal Age | 0.99 | *0.93* | 0.89 | 0.97 | 0.96 | 0.92 | 0.88 | 0.96 | 0.97 | 0.78 | 0.7 | 0.87 | 0.99 | *0.93* | 0.89 | 0.97 |
| Ipsi ACL reinjury | 0.92 | 0.93 | 0.91 | 0.95 | 0.95 | 0.93 | 0.96 | 0.9 | 0.92 | 0.86 | 0.95 | 0.77 | 0.96 | **0.94** | 0.96 | 0.92 |
| MCL Injury | 0.9 | **0.91** | 0.88 | 0.94 | 0.87 | 0.9 | 0.84 | 0.96 | 0.81 | 0.79 | 0.85 | 0.73 | 0.88 | 0.9 | 0.85 | 0.95 |
| LatMen Injury | 0.91 | **0.95** | 0.89 | 1.0 | 0.86 | 0.92 | 0.84 | 1.0 | 0.66 | 0.72 | 0.65 | 0.78 | 0.88 | 0.93 | 0.87 | 1.0 |
| MedMen Injury | 0.92 | 0.93 | 0.9 | 0.96 | 0.91 | 0.93 | 0.89 | 0.96 | 0.81 | 0.82 | 0.81 | 0.82 | 0.92 | **0.94** | 0.89 | 0.99 |
| PCL Injury | 0.97 | **0.98** | 0.97 | 1.0 | 0.98 | 0.94 | 0.98 | 0.9 | 0.88 | 0.75 | 0.89 | 0.6 | 0.98 | 0.94 | 0.99 | 0.9 |
| PLC Injury | 0.9 | 0.86 | 0.92 | 0.79 | 0.93 | **0.88** | 0.95 | 0.81 | 0.8 | 0.76 | 0.83 | 0.69 | 0.92 | 0.87 | 0.95 | 0.79 |

**Fig 5:**
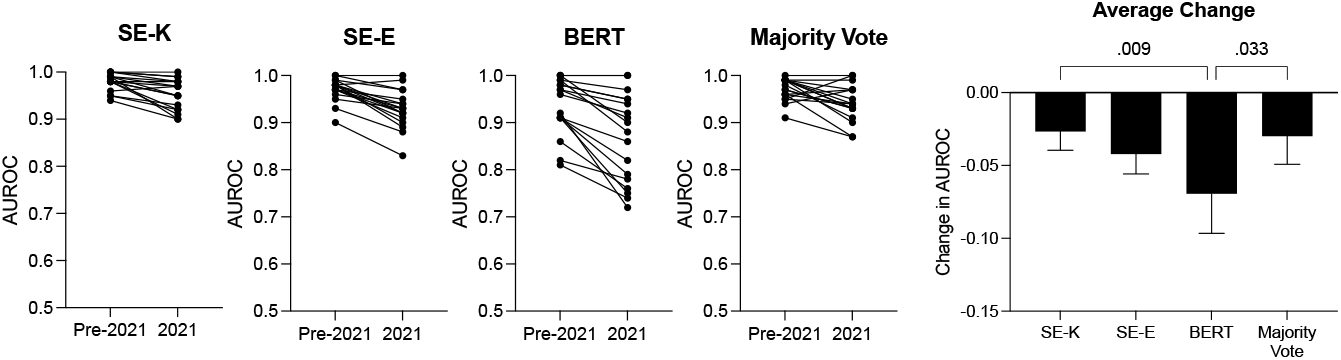
Comparison of results (AUROC) for original dataset (random test set drawn from 2000-2020) VS training on 2000-2020 data and predicting 2021 notes.

For the operative notes, there is no significant difference for AUROC when comparing the four approaches (null hypothesis of the Friedman test cannot be rejected P-value=0.47). The ranking of approaches for pre-operative notes is shown in Figure 6. In this set of results, SE-K and Majority vote are the best performing approaches and at the same time not significantly different from each other (P-value=0.76). Additionally, the results of both SE-K and SE-E are significantly better than BERT results (both P-values equal 0.02). Overall, SE-K and Majority vote results for 2021 data both have an average of 0.94 and std of 0.04 for AUROC while SE-E has an average of 0.93, std of 0.04 and BERT has 0.87 and 0.09 for average and std respectively.

**Fig 6:**
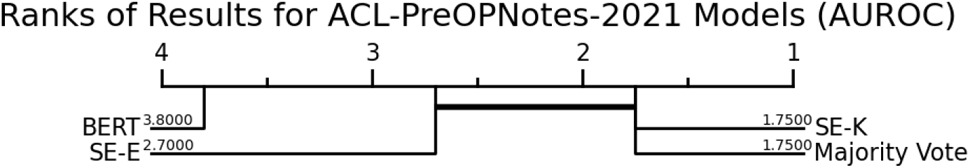
Ranking of different approaches based on AUROC in predicting the pre-operative variables from 2021 clinical notes. The horizontal lines indicate lack of significant difference between the 2 approach.

### 3.3 Efficiency

In comparing the computational efficacy of different approaches, Table 5 shows the average and standard deviation of elapsed time for each approach on different variables. For BERT, the times are reported on GPU while SE-K and SE-E are on CPU. Training BERT models on CPU takes more than 10 hours for each variable and is not a viable solution. SE-K has the fastest speed (at least 6 times faster on average, P-value<0.01). For pre-operative variables running times for SE-E and BERT are not statistically significant (P-value=0.14), however for operative variables, BERT (on GPU) is 2 times faster than SE-E (on CPU) on average (P-value=4e*-*7)

**Table 5:** Elapsed time for running different approaches in seconds. SE-K and SE-E were executed on CPU while BERT was executed on GPU.

|  | SE-K |  | SE-E |  | BERT(GPU) |  |
| --- | --- | --- | --- | --- | --- | --- |
|  | Mean | STD | MEAN | STD | MEAN | STD |
| OP Variables | 170.57 | 11.65 | 2740.10 | 366.88 | 1046.57 | 350.75 |
| Pre-OP Variables | 106.95 | 73.62 | 1748.01 | 1636.37 | 795.08 | 719.45 |

## 4 Discussion

In this paper, we assessed the ability of 2 proposed classification approaches SE-K and SE-E to extract information from unstructured clinical documentation. Overall, we showed that these approaches can be effectively used to build databases (e.g., registries) by extracting relevant information from unstructured clinical notes. The results showed superior performance of SE-K that is based on traditional NLP approaches (i.e., TF-IDF), compared to the state-of-the-art BERT approach in both in-sample and out-of-sample validation tasks. Also of note, the SE-K was able to extract these variables more efficiently than BERT, both in terms of time and resources (CPU vs GPU).

When validating these models on in-sample notes from 2000-2020, the ensemble method and SE-K method consistently outperformed BERT and SE-E. The fact that the ensemble method outperformed the other three models is not surprising, as other groups have found that combining different approaches leads to higher precision, recall, and accuracy than the individual methods alone [22, 23]. What is surprising, however, is the observation that SE-K, a model based on TF-IDF, consistently outperformed the more resource-intensive state-of-the-art BERT in extracting information from clinical notes, additionally in many cases had similar results to the majority vote which is usually the best performing model. Of note, TF-IDF has been used with much success recently in research involving clinical notes and disease classification. In their work, Dessi et al observed that their TF-IDF model with SVM classification outperformed word2vec[46], GloVe[47], and another word embedding algorithm, in identifying 16 different causes of morbidity from clinical notes [48]. They proposed that certain features made the classification strongly biased, allowing simpler approaches to more efficiently classify the different disease states. Similarly, Oh et al observed that TF-IDF, word2vec, and a combined hybrid approach all outperformed BERT in identifying literature involving drug-induced liver injury [22]. These results, as well as ours, suggest that BERT and similar embedding methods, as is, may not be the best candidates for clinical data, although they have dominated almost every other domain they have been applied to. This observation may be due to the format and nature of the provided text. Notably, this clinical documentation is often written with the use of a template, and many physicians use their own template, leading to little variation in content between unique notes. As observed in Figure 4, the clinical notes in this study cluster almost completely based on the primary surgeon (and note author). This signal based on the surgeon and author of the note could be so strong that it may complicate extracting other variables from the text. Notably, in keyword-based methods such as SE-K, the model ignores the words in templates, which may allow them to focus on the text unique to that specific note.

Additionally, we observed that the less resource-intensive SE-K and SE-E models outperformed BERT in the out-of-sample validation on the notes from 2021. In general, all models performed worse on the out-of-sample notes, possibly due in part to extrinsic factors such as a change in techniques, change in templates, or new surgeons. However, there was a significant decrease in performance with BERT compared to the other models. This suggests that the less intensive sentence extraction techniques may be more generalizable, and less likely to overfit. This is similar to the findings of Ezen-Can, who found that simpler models can achieve significantly better performance than BERT on smaller datasets and can be trained in much less time [49].

We also observed that the SE-K was able to run at faster speeds on the CPU, compared to BERT which required a GPU. In using sentence extraction methods for tasks such as those presented here, users can save a notable amount of time, and there is an additional financial and environmental benefit as well. BERT and other large language models have become increasingly popular in recent years, and there is evidence to suggest that making the models even larger can lead to further increases in downstream performance [50]. However, the computational and memory requirements of these large models prevent the widespread adoption for routine tasks. In their work, Strubell et al outline the sizable financial and environmental costs of training NLP pipelines and transformers using a GPU [51]. They equate the carbon footprint of training BERT on GPU to a trans-American flight. It is important for researchers to be aware of these costs at the onset of their study design phase, and our results as well as others suggest that less intensive methods are more than sufficient for extracting variables from unstructured clinical documentation.

Another advantage of SE-K and SE-E approaches compared to BERT is their interpretability. Extracting important keywords (Figure 3), and important sentences (Figure 1) from clinical notes, gives us the ability to better understand, and debug the model, and even the data itself, the classification task, and the labels.

Based on all the results, it is evident that SE-K is superior to SE-E. Given that SE-E is more like an extension of SE-K with doc2vec embeddings and it was proposed to improve SE-K performance, it begs the question that why is SE-E performing worse than SE-K. The answer could be one of two possibilities. Either doc2vec embeddings, similar to BERT embeddings, suffer from problems such as template effects. Or that doc2vec embeddings in general are not powerful enough to represent important sentences. Although answering this question requires more in-depth experiments, the available results favor the second answer. In SE-E doc2vec embeddings are only extracted for sentences that were identified as important by the model, in most cases these sentences do not have the template effect as much as other sections in the note. Similarly, we have seen improved results when feeding BERT models with important sentences instead of full clinical notes. Hence a possible direction for future work would be to replace doc2vec embeddings with BERT embeddings in SE-E and compare the results.

This study is not without limitations. Many of the limitations may be attributed to the nature of the training data. The language used in clinical notes lacks standardization and can be ambiguous, with the same word or phrase having different meanings in different contexts. This can make it challenging for NLP algorithms to accurately identify the relevant information in the notes. Additionally, clinical notes may not always be complete or accurate, and NLP algorithms may not be able to correctly interpret or extract information from incomplete or incorrect notes. Importantly, the models developed in this study may not be sufficiently generalizable to other institutions. Finally, there are ethical considerations in using these models, as there are no standard methodologies to respect the confidentiality of patients’ medical information when using these models to extract information.

## 5 Conclusion

In conclusion, this study shows that our proposed approach, SE-K, can be effectively used to extract relevant variables from clinic notes in order to build large-scale registries. We see consistently better performance with SE-K, compared to the more resource-intensive BERT. Additionally, this approach also provides interpretability which gives a better understanding of the model and the data. By building an ACL reconstruction registry using NLP, we can gain a better understanding of the outcomes of this surgery and identify potential areas for improvement. This can ultimately lead to better patient outcomes with a focus on individualized patient care.

## Data Availability

The data used in this study cannot be publicly shared due to privacy concerns. Data can be access upon reasonable request pending approvals from Boston Children's Hospital.

